# The predictive significance of prognostic nutritional index and serum albumin/globulin ratio on the overall survival of penile cancer patients undergoing penectomy

**DOI:** 10.1101/2022.03.29.22273150

**Authors:** Wei-Jie Song, Ni Chu-Jie Li, Jun Gao, Zhi-Peng Xu, Jian-Ye Liu, Zhi Long, Le-Ye He

## Abstract

**Objective:** To assess the value of using the prognostic nutritional index (PNI) and serum albumin/globulin ratio (AGR) in predicting the overall survival (OS) of patients with penile cancer (PC) undergoing penectomy.

**Materials and methods:** A retrospective analysis of 123 patients who were admitted to our hospital due to PC from April 2010 to September 2021 and underwent penectomy were included in the study. The optimal cut-off value of PNI and AGR was determined by receiver operating characteristic curve analysis. Kaplan-Meier analysis and the Cox proportional hazard model were used to evaluate the correlation between PNI, AGR, and OS in patients with PC.

**Results:** The best cut-off values of PNI and AGR were set to 49.03 (95% confidence interval 0.705-0.888, Youden index=0.517, sensitivity=57.9%, specificity=93.7%, P<0.001) and 1.28 (95% confidence interval 0.610-0.860, Youden index=0.404, sensitivity=84.1%, specificity=56.2%, P=0.003). Kaplan-Meier analysis showed that the OS of the patients in the high PNI group and the high AGR group was significantly higher than that of the patients in the low PNI group and the low AGR group (P<0.001). Univariate analysis showed that patient age, clinical N stage, pathological stage, PNI, and SII are all predictors of OS in patients with PC (P<0.05). Multivariate analysis showed that pathological stage (P=0.005), PNI (P=0.021), and AGR (P=0.004) are independent prognostic factors for predicting OS in patients with PC undergoing penectomy.

**Conclusions:** Both PNI score and serum AGR are independent prognostic factors for predicting OS in patients with PC undergoing penectomy.

## Introduction

Penile cancer (PC) is a malignant tumor that originates from the head of the penis, the coronal sulcus, the inner plate mucosa of the foreskin, or the skin of the penis. Due to demographic differences in many countries, including ethnic groups, religions, and sanitary conditions, the incidence of PC in different parts of the world varies widely. The incidence of PC has been reported to be as high as 19:100,000 men in some underdeveloped Asian, African, and Latin American countries, while the incidence in developed regions, such as Europe and the United States, is only about 0.1-0.9:100,000 men [1]. With the continuous improvement of Chinese medical and health level, PC has now become a rare androgenic malignant tumor. PC is mainly treated with penis preservation; however, partial or total penectomy is still the best choice for treatment for late stage or deep infiltrating tumors in the head of the penis [2]. Prognostic assessments PC, are mainly based on histopathology and clinical staging, and there is a lack of effective clinical biomarkers [3]. Therefore, identifying novel clinical prognostic parameters to assist in predicting the clinical outcome of patients with PC after penectomy is critical.

## Patients and methods

### Patients

We retrospectively analyzed the clinical data of 123 patients who underwent penectomy (partial/total penectomy) in our hospital from April 2010 to September 2021. All patients were diagnosed with PC through clinical laboratory testing, imaging examinations, and biopsy of penile masses before surgery. The exclusion criteria included: 1. Incomplete clinical data or follow-up data; 2. Patients suffering from diseases that affect systemic nutrition or immune status, such as autoimmune diseases; 3. Patients with blood infections or blood diseases, such as leukemia; 4. Patients taking any immunosuppressive drugs or any nutritional supplements before surgery; 5. Patients with other tumor diseases or with lymph node or distant metastasis before surgery.

### Data Collection

All patients included in the study underwent routine blood work and liver and kidney function assessments one week before surgery. The definitions of prognostic nutritional index (PNI), albumin/globulin ratio (AGR), systemic immune-inflammation index (SII), neutrophil-lymphocyte ratio (NLR), and platelet-lymphocyte ratio (PLR) were shown as follows: PNI = 1 × serum albumin (g/L) + 5 × lymphocyte count (109/L); AGR=serum albumin/ (total serum protein-serum albumin); SII=platelet × neutrophil/lymphocyte count; NLR = neutrophil/lymphocyte count; and PLR=platelet/lymphocyte count. The collected clinical pathological data included the age of the patient, the scope of penile resection, necrosis, clinical T and N grades, and pathological types. The clinical staging of tumors was determined according to the Tumor, Lymph Node, and Metastasis (TNM) staging system of the Joint Committee on Cancer of the United States (7th Edition) [4].

### Follow-up

Follow-up information was obtained by searching for outpatient review information and telephone follow-up with patients. The follow-up time was defined as the date of diagnosis of PC and penectomy to September 31, 2021. Follow-up occurred every four months for the first two years after surgery, and every six months from the third year after surgery onward. The follow-up included a physical examination, functional assessment, blood biochemical test, and a thorough examination of patients to determine any possibility of recurrence or metastasis.

### Statistical Analysis

All data were processed using SPSS v25.0 statistical software. The highest value of the Youden index was calculated using the receiver operating characteristic (ROC) curve as the best cut-off value of PNI, AGR, SII, PLR, NLR, and grouped separately. When the measurement data conformed to a normal distribution, mean ± standard deviation is used. The measurement data that did not conform to a normal distribution were expressed as the median (range). Kaplan-Meier survival analysis and log-rank test were used to compare the effects of different groups on overall survival (OS) in patients with PC undergoing penectomy. The variables with significant differences in univariate analysis (P<0.05) were included in the Cox proportional hazards regression model for multivariate survival analysis, and the hazard ratio (HR) and 95% confidence interval (CI) were estimated. Differences with a P<0.05 were considered to be statistically significant.

## Results

### 1. Patients’ information

A total of 123 patients with PC who underwent penectomy were included in this study, of which 101 (82.1%) underwent partial penectomy and 22 (17.9%) underwent total penectomy. The median follow-up time was 58 months. The age of the patients ranged from 29 to 90 years old, with a median age of 59 years. There were 95 cases (77.2%) with T<2 in clinical stage, and 28 cases (22.8%) with T≥2. In clinical stage, there were 115 cases (93.5%) with N=0, and 8 cases (6.5%) with N>0. Pathological classification showed that there were 12 cases (9.8%) of carcinoma in situ and 111 cases (90.2%) of squamous cell carcinoma. The median values of preoperative PNI, AGR, SII, PLR, and NLR were 49.30, 1.51, 464.53, 118.52, and 2.33, respectively. Table 1 summarizes the clinicopathological characteristics of the patients in this study.

### 2. The best cut-off values of PNI, AGR, SII and PLR before treatment

All 123 patients in the study were followed up, and the median follow-up time was 58.0 months, during which 16 (13.0%) participants died. The cut-off values for PNI, AGR, SII and PLR were determined by the ROC curve, and the optimal cut-off values were set at 49.03, 1.28, 636.99, and 121.95, respectively. The Area Under the Curve (AUC) of the PNI was 0.797 (95% CI: 0.705-0.888, Youden index=0.517, sensitivity=57.9%, specificity=93.7%, P<0.001). The AUC of the AGR was 0.735 (95% Cl: 0.610-0.860, Youden index=0.404, sensitivity=84.1%, specificity=56.2%, P=0.003). The AUC of the SII was 0.657 (95% Cl: 0.505-0.808, Youden index=0.326, sensitivity=62.5%, specificity=70.1%, P=0.044). The AUC of the PLR was 0.654 (95% Cl: 0.521-0.787, Youden index=0.320, sensitivity=75.0%, specificity=57.0%, P=0.047) (Figure 1).

**FIGURE 1.**
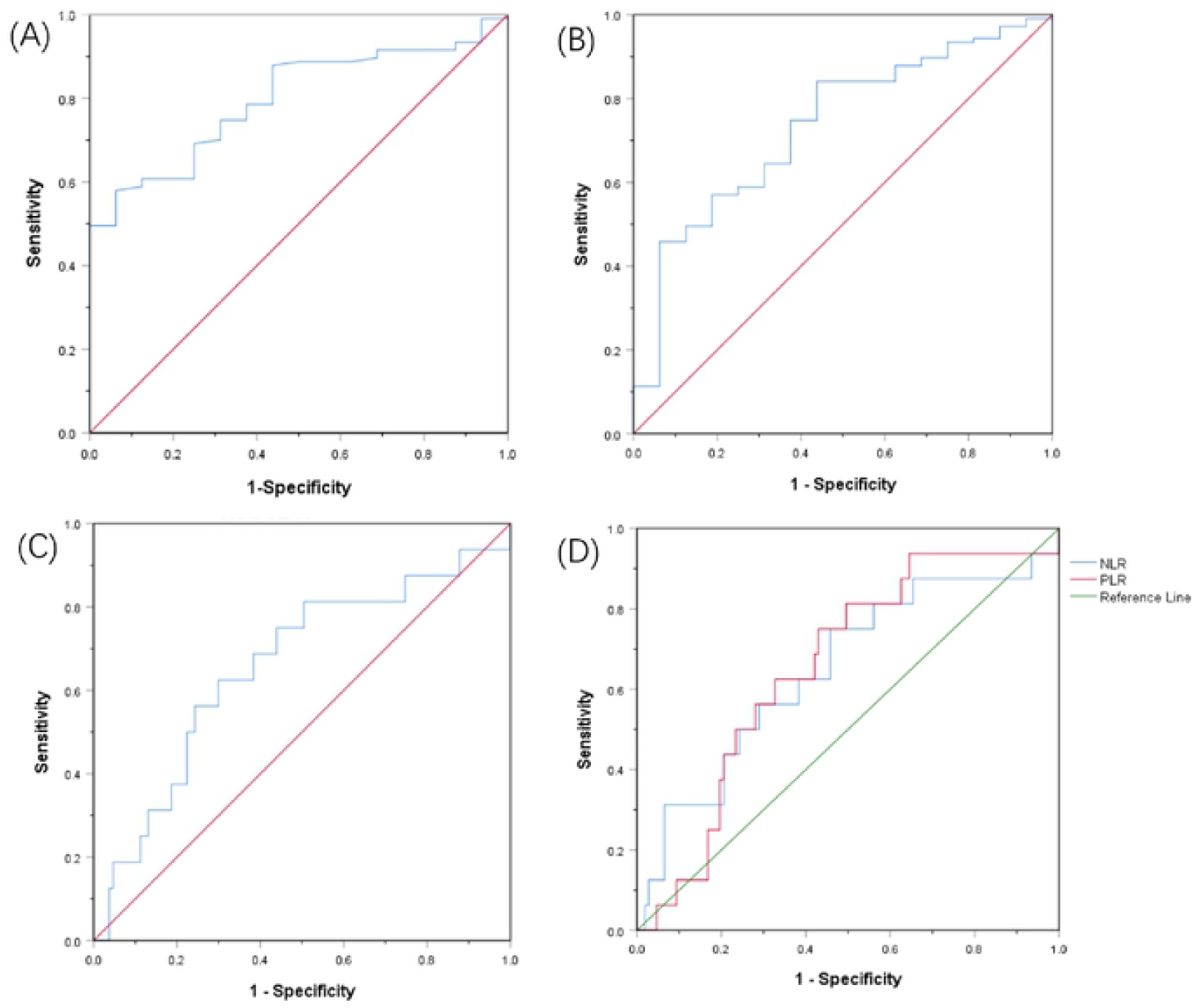
Receiver operating characteristic curves for pretreatment (A) PNI, (B) AGR, (C) SIl, (D) RLR and NLR based on OS.

### 3. Comparison of clinical data of patients in different groups

The optimal cut-off values of PNI, AGR, SII, and PLR were used as the standard, and patients below the cut-off value were defined as the low-level group, while those higher than or equal to the cut-off value were defined as the high-level group. Because the maximum area under the curve when NLR=2.34 is 0.648 (95%Cl 0.493-0.803, Youden index=0.292, sensitivity=75.0%, specificity=54.2%, P=0.057), P>0.05 has no statistical significance. Therefore, it is not used as a standard for grouping. The clinical N stage of patients in the low AGR group was worse than that in the high AGR group (P=0.039, Table 2). In addition, compared with the low SII group, the high SII group retained less penile tissue. (P=0.027, Table 2).

### 4. Kaplan-Meier Survival Analysis

Kaplan-Meier survival analysis showed that the median OS of patients with PNI<49.03 was 100.4 months, which was lower than the median OS of patients with PNI≥49.03 of 135.8months (P<0.001). The median OS of patients with AGR ≥ 1.28 before surgery was 128.2 months, which was significantly higher than the median OS of 75.7 months for patients with AGR <1.28 (P<0.001). The median OS of patients with SII≥636.99 before surgery was 10.5 months, which was significantly lower than the median OS of patients with SII<636.99 of 128.0 months (P=0.010). The median OS of patients with PLR≥121.95 before surgery was 104.2 months, which was significantly lower than the median OS of patients with PLR<121.95 of 129.9 months (P=0.012) (Figure 2).

**FIGURE 2.**
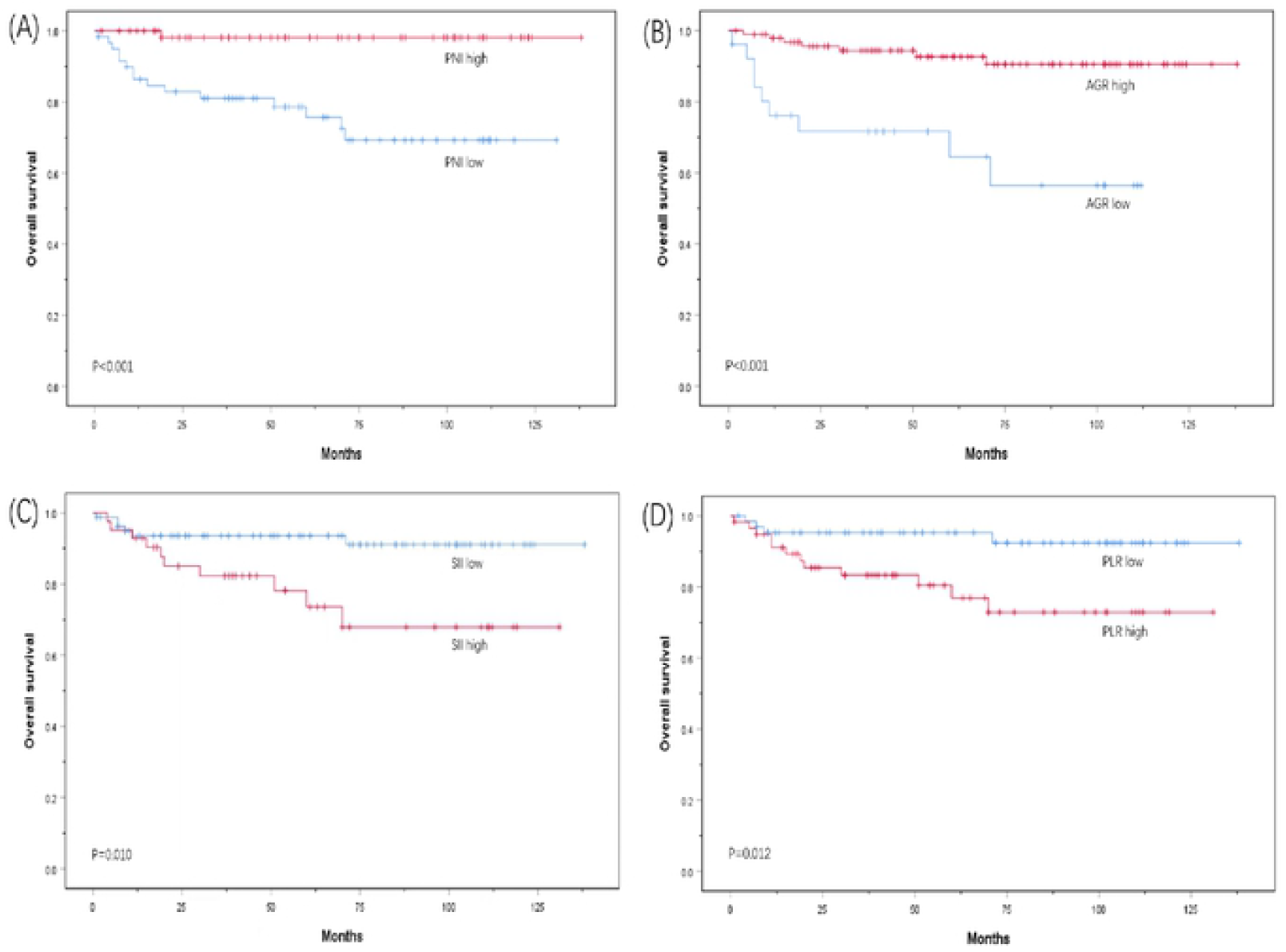
Kaplan-Meier survival curves for OS according to (A) PNI, (B) AGR, (C) Sll and (D) PLR in PC patients.

### 5. Univariate and multivariate analysis

The results of the univariate analysis showed that clinical N staging, pathological classification, preoperative PNI, AGR, SII, PLR, and OS of patients with PC were statistically correlated (all P<0.05, Table 3). Multivariate analysis revealed that preoperative PNI (risk rate [HR] = 0.057; 95% CI: 0.005–0.645; P = 0.021; Table 3) and AGR (risk rate [HR] = 0.159; 95% CI: 0.046–0.554); P = 0.004; Table 3) are both independent prognostic factors for OS in patients with PC treated with penectomy.

## Discussion

PC is a relatively rare androgenic malignant tumor, which can seriously affect the quality of life of men. Penectomy is one of the main treatment methods for PC, and postoperative OS is the most important prognostic indicator that clinicians and patients pay attention to. There is still a lack of specific prognostic biomarkers for PC, so it is very important to explore reliable clinical markers for the diagnosis and treatment of PC. In this study, we evaluated the prognostic value of PNI score and serum AGR in predicting OS in patients with PC. Both PNI score and serum AGR can reflect the systemic nutrition and inflammation status of tumor patients, and both are non-invasive biomarkers.

The latest research shows that systemic malnutrition and inflammation are closely related to the poor prognosis of various malignant tumors [5,6]. Thus far, a variety of proteins have been found in human plasma, among which albumin accounts for more than half [7]. Serum albumin (ALB), a central water-soluble protein produced by the liver, maintains the osmotic pressure of capillaries and eliminates free radicals in the blood. It is also commonly used in clinical practice to assess the nutritional status of patients [8]. Malnutrition can lead to delayed postoperative healing and decreased collagen synthesis, thereby damaging the body’s immune system [9]. At the same time, malnutrition can lead to increased mortality after surgery and reduce the resistance of patients to malignant tumors [10]. A study on ALB and cancer found that the serum ALB level of cancer subjects was significantly lower than that of non-cancer subjects [11]. ALB is also closely related to the prognosis of a variety of cancers, and can be used as a predictor of recurrence, metastasis, and death of a variety of malignant tumors [12-14]. In a study of a physical examination crowd, the correlation between low levels of ALB and tumor morbidity and mortality was also found [15]. In addition to being a nutritional marker, ALB is also an inflammatory response index [16]. Various pro-inflammatory cytokines can regulate the synthesis of albumin in hepatocytes as part of the tumor inflammatory response and, at the same time, can promote tumor growth and metastasis, destroy the host’s immune response, and promote drug resistance [17]. Numerous studies have confirmed that markers of chronic inflammation play a key role in cancer recurrence, metastasis, and progression, and are closely related to the OS of a variety of malignant tumors [18-20].

The PNI index is calculated from ALB and lymphocyte count and can reflect the body’s nutritional and inflammatory state to a certain extent. Lymphocytes also play a very important role in the immune surveillance of malignant tumors. They can inhibit the proliferation and metastasis of malignant tumors by promoting the production of cytokines and the apoptosis of cytotoxic cells [21]. At present, researchers have studied the role of the PNI index in the prognosis of various malignant tumors. The total tumor recurrence rate and OS of patients in the higher PNI score group were higher than those in the lower PNI score group. The PNI index has been found to be an independent prognostic factor for a variety of malignant tumors, but none of these reports assessed the prognosis of PC [22-24].

Serum AGR is calculated by the ratio of ALB to serum globulin and can reflect the patient’s nutritional and inflammatory state. Serum globulin is calculated by subtracting ALB from total serum protein. Therefore, it contains a variety of pro-inflammatory proteins, such as C-reactive protein (CRP), immunoglobulin (Igs) and complement components. Studies have shown that elevated levels of CRP before surgery can lead to poor cancer prognosis [25]. Studies of patients with lung cancer have also found that a higher serum globulin level was associated with a lower survival rate [26]. Additionally, studies of patients with colorectal cancer have found that high complement and IgA levels are also poor prognosis indicators [27]. Serum AGR has also been confirmed as an independent prognostic factor for a variety of malignant tumors [28-30].

In this study, we found that both PNI score and serum AGR can be used as independent prognostic factors for predicting OS in patients with PC undergoing penectomy. Higher PNI scores and higher AGR levels are associated with longer OS for patients with PC. We confirmed the importance of preoperative nutritional support and inflammation control for the prognosis of patients with PC undergoing penectomy. Additionally, we found that poor clinical staging and higher levels of PLR and SII are related to the poor prognosis of patients with PC. PLR and SII are also commonly used clinical nutrition and inflammation markers, once again corroborating the results of this study. However, because this was a single-center study, the sample size was relatively small. Therefore, whether preoperative PNI score and AGR are valid to prognosis indicators for patients with PC requires further investigation and verification.

## Data Availability

All relevant data are within the manuscript and its Supporting Information files.

## Disclosure

### Ethical consideration

Registry and the Registration No. of the study/trail: N/A. Animal Studies: N/A.

### Ethics approval

Approve by Institutional Review Board (IRB) of The Third Xiangya Hospital of Central South University (Grant No. 21154).

### Consent for publication

All authors consent for the publication of the article.

## Acknowledgements

All work was completed at the Department of Urology, Central South University, The Third Xiangya Hospital. The authors thank AiMi Academic Services (www.aimieditor.com) for the English language editing and review services.

## Authors’ Contributions

WS and NC initiated the research question and supervised all aspects of the study. JG and ZX contributed to data acquisition. JL supervised the statistical procedures of the manuscript. WS and NC wrote the first version of the manuscript. ZL and LH reviewed this article.

